# Artificial Intelligence in laryngeal endoscopy: Systematic Review and Meta-Analysis

**DOI:** 10.1101/2022.01.16.22269346

**Authors:** Michał Żurek, Anna Rzepakowska, Kamil Jasak, Kazimierz Niemczyk

## Abstract

**Background:** Early and proper diagnosis of laryngeal lesions is necessary to begin treatment of the patient as soon as possible with the possibility of preserve organ functions. Imaging examinations are oft aided by artificial intelligence (AI) to improve quality and facilitate appropriate diagnosis. The aim of the study is to investigate of the diagnostic utility of AI in laryngeal endoscopy.

**Methods:** Five electronic databases (PubMed, Embase, Cochrane, Scopus, Web of Science) were searched for studies published before October 15, 2021 implementing artificial intelligence (AI) enhanced models assessing images of laryngeal lesions taken during laryngeal endoscopy. Outcomes were analyzed in terms of accuracy, sensitivity and specificity.

**Results:** All 13 included studies presented overall low risk of bias. The overall accuracy of AI models was very high (from 0.806 to 0.997) and the number of images used to build and evaluate the models ranged from 120 to 24,667. The accuracy was significantly higher in studies using larger database. The pooled sensitivity and specificity for identification of healthy laryngeal tissue (8 studies) was 0.91 (95% CI: 0.83-0.98) and 0.97 (95% CI: 0.96-0.99), respectively. The same values for differentiation between benign and malignant lesions (7 studies) were 0.91 (95% CI: 0.86-0.96) and 0.95 (95% CI: 0.90-0.99), respectively. The analysis was extended to a comparison of sensitivity and specificity of AI models assessing Narrow Band Imaging (3 studies) and white light endoscopy images (4 studies). The results were similar for both methods, no subgroup effect was revealed (p = 0.406 for sensitivity and p = 0.817 for specificity).

**Conclusions:** In assessing images of laryngeal lesions, AI demonstrates extraordinarily high accuracy, sensitivity, and specificity. AI enhanced diagnostic tools should be introduced into everyday clinical work. The performance of AI diagnoses increases efficacy with the size of the image database when using similar standards for evaluating images. The multicentre cooperation should concentrate on creation of huge database of laryngeal lesions images and implement their sharing, which allows building AI modes with the best performance, based on vast amount of images for learning and testing.

## 1. Introduction

The spectrum of laryngeal pathologies is very wide, and every level of larynx may be involved in neoplastic, pre-neoplastic or non-neoplastic process, however the majority of changes localize in glottic part. Prior to laryngeal cancer, cellular changes begin with epithelial hyperplasia, then develop into dysplasia, squamous cell carcinoma in situ and eventually into invasive cancer [1, 2]. Potentially, 6% to 22% of premalignant lesions will develop into malignancies, and the transformation rate depends on severity of the precancerous lesions [2]. The aim of modern diagnostics is the proper assessment of lesions in the larynx with the lowest possible invasiveness of the examination. First, it is necessary to distinguish malignant and potentially malignant lesions from benign ones. The benign vocal fold lesions classification includes nodules, polyps, cysts, fibrous masses, pseudocysts, and non-specific lesions [3].

It is crucial to perform early diagnosis and preoperative assessment in order to provide adequate and least invasive treatment to preserve organ functions [2]. Especially in case of laryngeal cancer and its precursor lesions, treatment process has a great influence on everyday basic functions such as breathing, swallowing and voice production. Current approach to laryngeal cancer put great impact on avoiding laryngectomy whenever possible in order to maintain the best quality of life [4-6]. Low advanced precancerous lesions usually require minimally invasive resection of the lesion. In case of high-grade dysplasia and carcinoma in situ, laser resection and ablation may be also necessary [7]. For malignancies, the treatment is more demanding and require resection with the margin of healthy tissue. Endoscopic intralaryngeal resection and open partial laryngectomy are used to achieve complete excision while maintaining the larynx in patients with early-stage malignancies (T1 and T2). Total laryngectomy or chemoradiation therapy has become the primary treatment for patients with advanced cancers (T3 and T4) [4].

Many tools are used in the diagnosis of laryngeal lesions at different stages of advancement, including indirect and direct laryngoscopy, ultrasound, computer tomography, and magnetic resonance imaging [2]. Each of the methods has its advantages and limitations, which affects their usefulness. The diagnosis of laryngeal lesions begins mostly with indirect laryngoscopy, preferably with the endoscopy equipment [8]. The speed, easiness of performance, low cost, and high efficiency of endoscopy have made it a key diagnostic tool. Various facilities have been introduced to improve the sensitivity and specificity of this examination. The original white light endoscopy (WLE) imaging has certain limitations, and the images of different dysplasia stages and early cancer may have the same appearance. WLE is not so precise in distinguishing mucosal differences, especially when assessing dysplastic and cancerous lesions at early stages [6]. Because of certain limitations of classical WLE, some image-enhanced endoscopy techniques such as autofluorescence, contact endoscopy, narrow band imaging (NBI) and Storz Professional Image Enhancement System (SPIES) have been developed. Nowadays, there is an increase in usage of these enhanced endoscopy techniques observed in everyday clinical practice, especially among patients with laryngeal pathologies [9]. Particularly NBI and SPIES techniques has been shown to be more accurate in diagnosing laryngeal dysplasia comparing to WLE alone [10]. According to Stanikova et al. it was proved that NBI, as well as SPIES has similar potential in differentiation between low-risk and high-risk lesions. The sensitivity was 83% and 86% respectively for NBI and SPIES, and specificity of 98% was demonstrated for NBI, and 96% for SPIES [11]. It should be emphasized that directed biopsy and histopathology remains the gold standard for final diagnosing of laryngeal lesions. However, biopsy is a mentally and physically demanding procedure for the patient and may cause vocal fold or laryngeal dysfunction [5, 12], therefore less invasive diagnostic methods of sensitivity and specificity closest to histopathology results are still searched.

One of the crucial problems related with introducing new diagnostic tool is the learning process. The relationship between efficiency and experience is not a linear dependence. The learning speed changes depending on the level of the examined person [13]. In order to avoid limitations in the access to the recent diagnostic methods due to the lack of experience of young doctors, many computer facilities supporting the assessment of lesions are currently implemented. One of such tools is artificial intelligence (AI). Since its beginning in the fifties, AI has evolved dramatically. Nowadays, AI may precipitate diagnosis, improve its accuracy and induce positive impact on efficiency in clinical practice. The fact that some subclasses of AI allow machines to learn how to use gathered information and make decisions independently is very promising. The AI can analyze an input image to recognize patterns and create specific filters in order to compute the final outcome [14]. Introducing AI into diagnostic process and medical images analysis contributed to better precision, replicability, and efficiency in making diagnosis. In 2017 Arterys became the first U.S. Food and Drug Administration approved clinical application in healthcare, based on cloud-storage data [14]. CardioAI was the first Arterys product, used in analysis of magnetic resonance heart images. Since then, the application has been developed to analyze also liver and lung imaging, chest x-ray and bone x-ray images, and head CT images without contrast. Applications based on AI widely found use in gastroenterology, where two systems are eligible for usage now:

- ENDOANGEL (Wuhan, China), a system developed in 2019, which is able to objectively assess bowel preparation for endoscopy;
- GI Genius (Minneapolis, USA), is a tool used during endoscopy, which help endoscopist to identify colorectal polyps by showing a visual marker on a screen at the time of examination.

The advantages of AI-enhanced systems have been proven many times, an example is the study of Repici et al., where a 14% increase in adenoma detection rate was noted using AI system [15].

The purpose of this meta-analysis is to evaluate the efficacy and clinical utility of AI in the assessment of laryngeal lesions based on endoscopic images. The objective will be achieved by analyzing the ability of AI to evaluate selected laryngeal lesions based on accuracy, sensitivity and specificity.

## 2. Materials and Methods

### 2.1. Search Methods, Types of Studies, and Participants

A systematic review of the literature was undertaken to investigate the diagnostic utility of AI in laryngeal endoscopy. To report the results as recommended, PRISMA guidelines [16] were followed. PICO framework model [17] was used to describe search strategy (Table 1). The search was conducted through five publication databases (PubMed, Embase, Cochrane, Scopus and Web of Science) by two independent scientists (MŻ and KJ), what allowed to retrieve two additional articles. Search strategies used in the systematic review are presented in Supplementary Materials (Table S1). Publications available until October 15, 2021 were included.

**Table 1.** Population, Intervention, Comparison, Outcome (PICO).

| PICOS framework |  |
| --- | --- |
| Population | Patients (without any age limit) who underwent laryngeal endoscopic examination |
| Intervention | Evaluation of endoscopy images by AI |
| Comparison | Histopathology or specialist assessment |
| Outcome | Classification of laryngeal lesions |

All patients who underwent laryngeal endoscopic examination were included in the study. Randomized controlled trials as well as retrospective and prospective cross-sectional studies, including case-control and cohort type accuracy studies were subsumed. Animal or in vitro studies, publications not written in English, case reports, reviews or systematic literature reviews, editorials and opinion pieces, meta-analysis, and conference abstracts were excluded.

### 2.2. Index Tests and Target Conditions

Studies that examined the sensitivity, specificity or accuracy of AI classifying laryngeal lesions based on endoscopic images were eligible. The reference standard were histopathology results or ENT specialist assessment.

### 2.3. Data Collection and Analysis / Selection process

Study selection was divided into three phases. First phase was the removing of duplicated results in EndNote 20 software (Clarivate Analytics, Philadelphia, PA, USA). Second phase was screening and filtering titles and abstracts of scientific papers against inclusion and exclusion criteria. The first and second phases were realized by two reviewers (MŻ and KJ). During the third phase independent reviewer (AR) evaluated the research’ full-text manuscripts for eligibility, noting the reasons for exclusions. Any inconsistencies between the reviewers were settled through conversation, until agreement was reached. A PRISMA flowchart [16] summarizing the results of data collection and analysis was created. The review protocol was registered with the International Prospective Register of Systematic Reviews (PROSPERO, CRD42021282843).

### 2.4. Risk of Bias Assessment

The quality of the studies was evaluated by three reviewers (MŻ, AR, KJ) independently using a quality assessment tool for diagnostic accuracy studies (QUADAS-2) [18]. The QUADAS-2 tool is divided into four primary domains: patient selection, index test, reference standard, flow of patients through the study and timing of the index tests and reference standard (flow and timing). According to the authors recommendations of QUADAS, questions should be review-specific tailored. Due to the specific nature of the assessed studies, the domain “Patient selection” was replaced by “Materials selection”. Furthermore, additional questions were added to each domain in QUADAS tool, and some of the original questions were omitted. The tailored QUADAS tool is presented in the supplementary table (Table S2). Based on the results of bias assessment clustered bar graphs were prepared.

### 2.5. Statistical Analysis and Data Synthesis

The aim of the study was to assess the clinical usefulness of AI in the laryngeal endoscopy, therefore the study focused on four main aspects:

1. Analysis of overall accuracy of AI in assessing laryngeal lesions;
2. The ability of AI to identify healthy tissue;
3. The ability of AI to differentiate benign lesions from premalignant and malignant ones;
4. Analysis of diagnostic performance of AI using NBI and WLE images.

Before proceeding to the comparative analysis of the selected studies, it was necessary to standardize the terminology of laryngeal lesions across the studies. Most of the authors used classification on heathy tissue, benign, precancerous and malignant lesions, however in some papers clinical terms for changes were applied. In these researches the cysts, nodules, polyps, Reinke’s edema, webs, sulcus vocalis and laryngitis were included into benign lesions. Keratosis, leukoplakia, dysplasia mild and severe and papillomatosis were considered as precancerous one. The same inhomogeneity was revealed for vascular pattern description of the involved laryngeal mucosa in NBI endoscopy. For consistency, it was decided to transform the nomenclature of vascularization in accordance with the most widespread classification of Ni classification. [19]. Raw data were extracted from each involved study in the form of a 2 × 2 table, including the numbers of true positives (TP), false positives (FP), true negatives (TN), and false negatives (FN). A summary of the data collected and the terminology used are presented in supplementary materials (Table S3 and S5).

A meta-analysis of diagnostic accuracy of raw data was conducted using R “meta” package version 5.0-1, “metafor” package version 3.0-2 and “nsROC” package version 1.1 (R version 4.0.2, R Foundation for Statistical Computing, Vienna, Austria). The forest plots and receiver operating characteristic (ROC) curves were performed to depict the relationship between individual and summarized values of specificity and sensitivity. T^2^ and I^2^ statistics were used to evaluate the study heterogeneity. To assess the heterogeneity between subgroups the test for subgroup differences was used. Sensitivity and specificity analyses using the random-effects model were conducted for both analyzed fields. Statistics with p-value under 0.05 were considered significant.

## 3. Results

### 3.1. Results of the Search

Based on the literature search on 15 October 2021 a total of 895 publications from five databases (PubMed, Embase, Cochrane, Scopus, Web of Science) were identified. There were 139 duplicate records, after removing 756 publications remained, which were screened by title and abstract. This led to the exclusion of 728 publications. The confrontation of the results of the literature review with another researcher resulted in retrieval of 2 additional records. So there were 30 publications included for full-text assessment. 17 publications were excluded thereafter. The systematic review included in total 13 papers evaluating the diagnostic accuracy of AI in laryngeal endoscopy [20-32], as shown in the PRISMA flow diagram (Figure 1) and summarized in supplementary Table S3.

**Figure 1.**
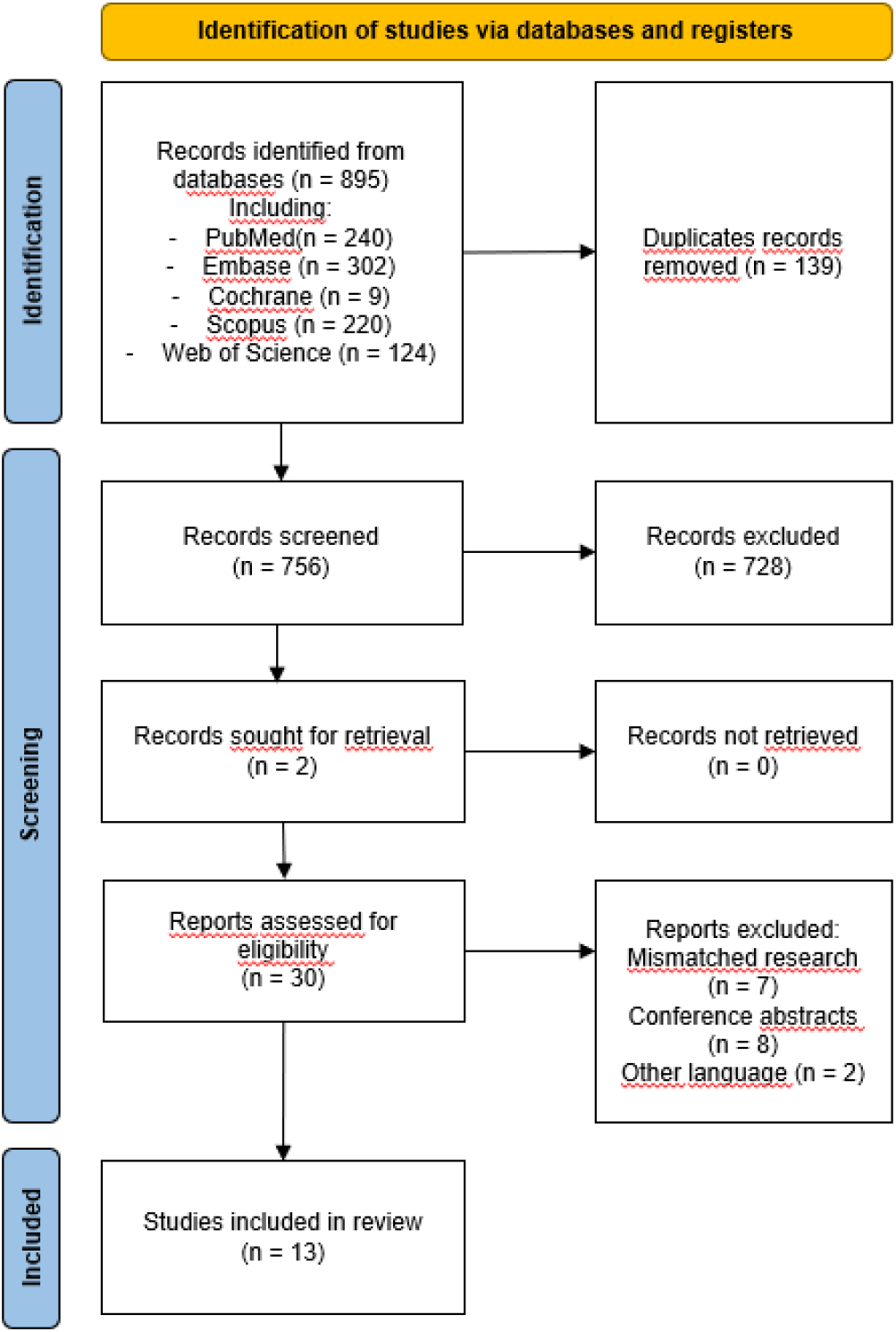
Flow diagram of the systematic review search.

All of the included studies were retrospective studies and used AI to assess images of laryngeal lesions. All neural networks assessed the character of the lesions on the basis of vascular patterns, shape and/or colour. 8 of them evaluated endoscopic images in white light and 5 using NBI method. The total amount of images used in individual study was widely varied, from 120 to 24,667. Also, the implicated by the authors pre-processing methods varied. In case of 8 studies, images of the entire vocal folds were used, while in 5 studies only selected fragments of their images were exploited. The methodology varied from manual selection of images and their classification to complex informatic methods allowing for extracting specific features of the images. 6 of the studies used pretrained convolutional neural network (CNN) to classify the lesions, others used support-vector machine (SVM), k-nearest neighbours (KNN) or random forest (RF) algorithm. The analysis evaluated the AI overall diagnostic accuracy in laryngeal endoscopic procedures [20-32]. Concerning different objectives of included studies, there were also performed subgroups analyses to verify the utility of AI in clinically specific diagnostic problems:

- Identification of healthy laryngeal tissue that included 8 studies [22, 23, 26-28, 30-32];
- Differentiation between benign and malignant laryngeal lesions that included 7 studies [23-27, 30, 32];
- Comparison of AI accuracy of white light endoscopy (4 studies) [23, 27, 30, 32] with NBI method (3 studies) [24-26].

### 3.2. Risk of Bias Assessment

The results of the QUADAS-2 bias and applicability evaluation are summarized in Figure 2, whereas Table S4 (Supplementary Materials) lists the specific bias scores for each of the seven categories for all research included. In a large number of the included research, the QUADAS-2 assessment revealed low risk of bias.

**Figure 2.**
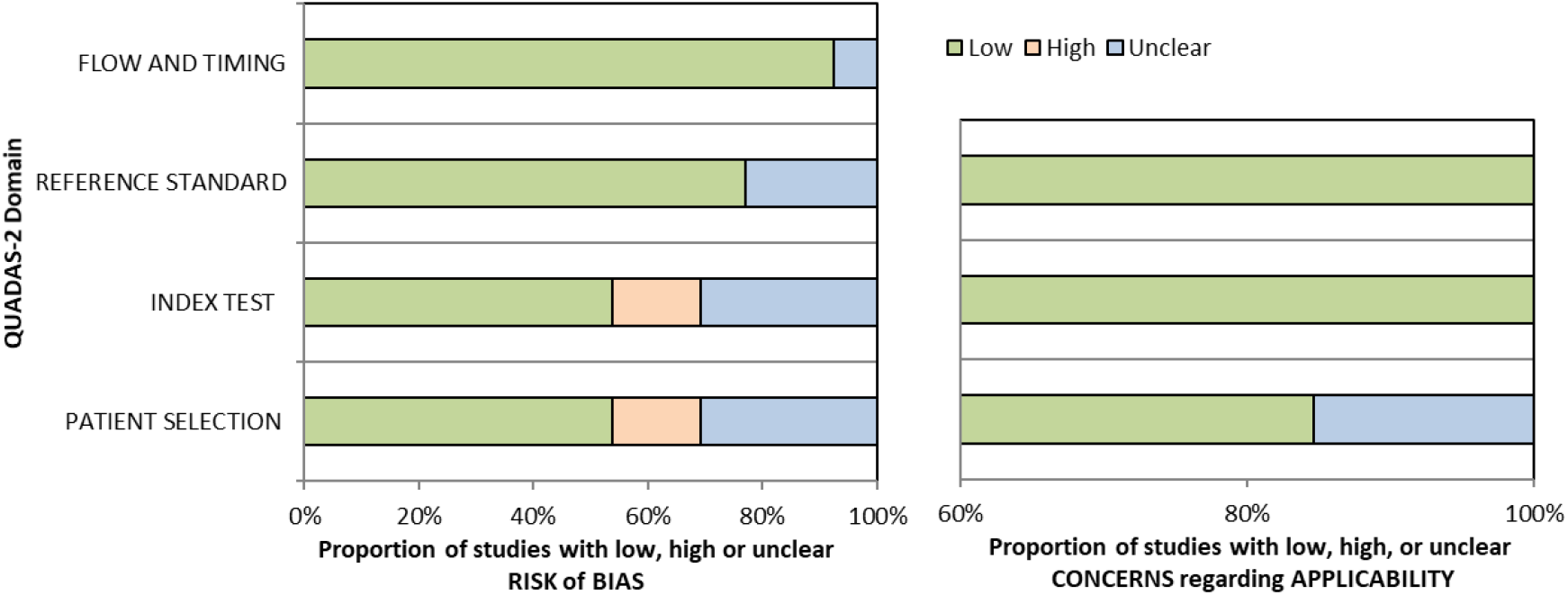
QUADAS-2 assessment of bias and applicability.

Bias in patient selection was low in 7, unclear in 4 and high in 2 studies. The material selection bias was difficult to assess because of the specific nature of the research. The selection process of patients was not always clear, and it was considered that the evaluation of the fragments of images may contribute to the reduced credibility of the research materials, which at the same time increases the risk of bias in the domain. The risk of bias in the index test and reference standard was high in 2 studies and 0 studies and unclear in 4 and 3 studies, respectively. The reason for this result was the lack of appropriate presentation of the results and the lack of confirmation of the diagnosis by histopathological examination. In one study, the flow and timing risk of bias was unclear, and for the rest of the included studies it was low. Moreover, considerable variation in terminology and pre-processing methods can lead to heterogeneity in all modalities. The risks of bias in all domains were mostly high and unclear in three studies [28-30]. The patient selection process in these studies has not been adequately described, and the reference standard is also not obvious. In two studies [20, 26] only fragments of images from patients with laryngeal carcinomas were used, which limits the randomness of the research group, therefore the bias in patient selection was considered high. The results in 3 studies [20, 21, 29] were not adequately presented, which limited their usefulness in this publication.

### 3.3. Diagnostic accuracy of AI in assessment of laryngeal lesions

The first part of the analysis includes the assessment of the accuracy of all included studies. Due to the variety of researches, in particular the objectives and number of research groups, the classic forest plot and ROC analysis is not recommended. The aim of this part is to indicate the potential of neural networks and their dependence on the number of images used. Accuracy of AI in assessment of laryngeal lesions differs between 0.806 to 0.997. Such high accuracy shows how valuable it is to introduce AI into everyday clinical work, regardless of the type of laryngeal lesion assessed.

Figure 3 shows the relationship between accuracy and number of images per research. The chart allow distinguishing and comparison of results of two types of researches with low and high amount of analysed images. In the first group of studies, a relatively small number of images beneath <2500 were analysed with AI and there was obtained a wide range of AI accuracy from 0.806 to 0.997 [20, 21, 24-26, 28-31]. There was also observed tendency of increasing accuracy with amount of applied pictures, however it must be also stressed that in each of these studies there were applied advanced and different pre-processing methods for images including: gaussian smoothing, investigation of texture-based global descriptors, calculating first-order statistics, specular reflections removal, region of interest (ROI) detection. A linear regression curve was determined for the first group. Its formula is as follows:

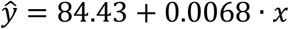

where y is accuracy and x is the number of images.

**Figure 3.**
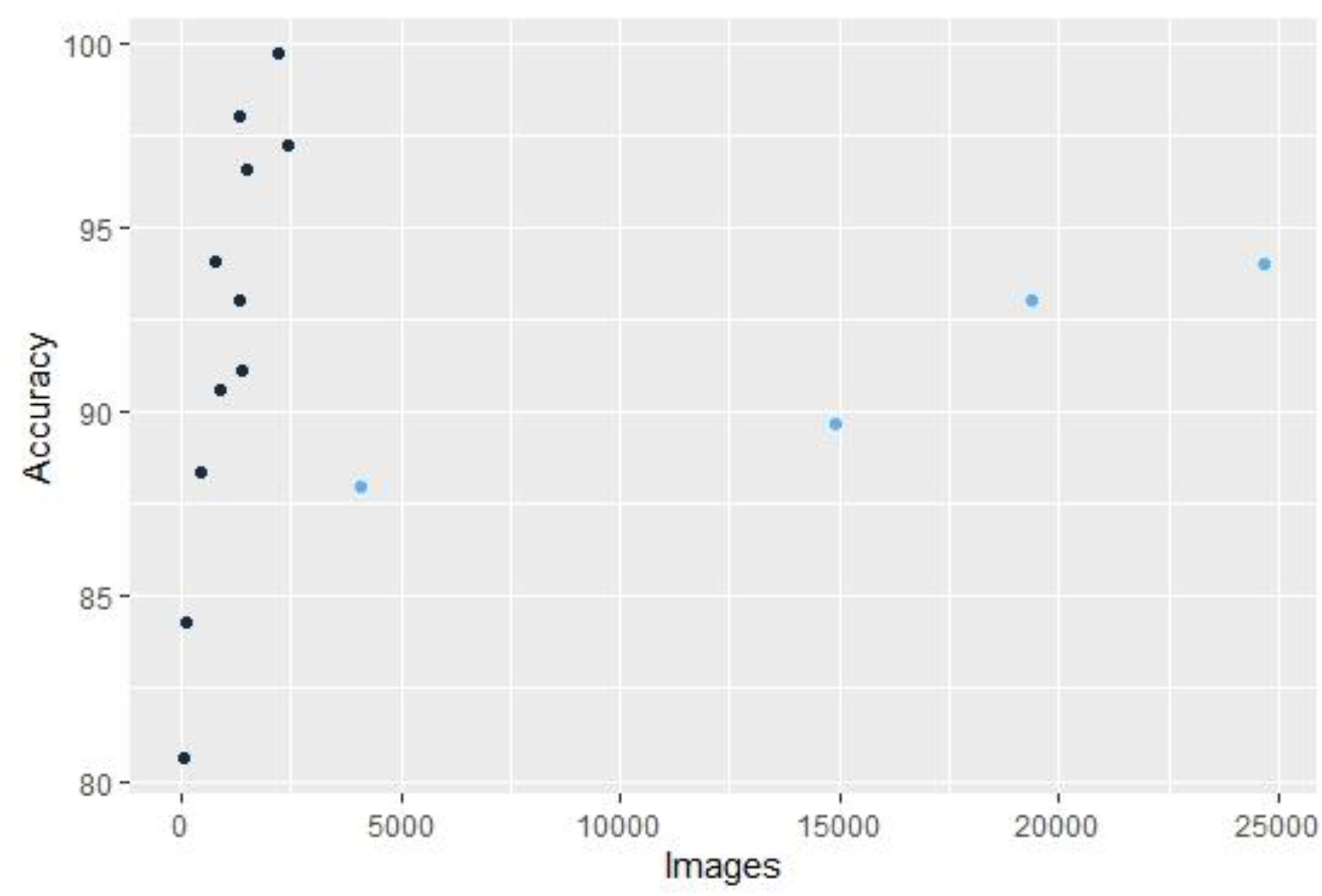
Dot plot of the accuracy of included studies (there are more dots than studies because some research analysed more than one classification of laryngeal lesions).

The assessment of the model fit is good R^2^ = 0.7649; p-value = 0.0004.

For the second group with images capacity exciding 2500 there was recognised an evident trend of increasing accuracy with the number of included images from 0.88 to 0.94 [22, 23, 27, 32]. This tendency cannot be confirmed statistically yet, due to only four performed on such scale studies so far, but it is worth noting that for these studies with huge material analysed, the pre-processing methods were very simple comparing to the first group and included only choosing images and detection of ROI. These comparison identifies two directions for future researches with the awareness of limitations of data preparation that should be unified and verified not to influenced the accuracy score.

Construction of a linear regression model for all studies would not be valid due to excessive differences in their methodology.

### 3.4. Diagnostic sensitivity and specificity for identification of normal tissue

The diagnostic performance of AI in identification of healthy laryngeal tissue during endoscopy is presented in supplementary materials (Table S5) and in Figure 3. The estimated mean sensitivity and specificity of the diagnosis of healthy tissue was 0.91 (95% CI: 0.83-0.98) and 0.97 (95% CI: 0.96-0.99), respectively. The area under ROC curve (AUC) was 0.994.

The between-study heterogeneity variance was estimated for polled sensitivity and specificity analysis and revealed a substantial difference between studies (Sensitivity: τ^2^ = 0.0063 (95%CI: 0.0021-0.316), I^2^ = 96.8% (95%CI: 95.1%-97.9%); Specificity: τ^2^ = 0.0001 (95%CI: 0.0001-0.0012), I^2^ = 82.7% (95%CI: 63.4%-91.8%); p-value for both analysis <0.0001).

### 3.5. Diagnostic sensitivity and specificity to distinguish between benign and malignant lesions

The next stage of the analysis concerned the assessment of the effectiveness of AI in distinguishing benign from malignant lesions in endoscopic examinations of the larynx. The diagnostic performance is presented in supplementary materials (Table S5) and in Figure 4. The estimated mean sensitivity and specificity of the differential diagnosis between benign and malignant lesions was 0.91 (95% CI: 0.86-0.96) and 0.95 (95% CI: 0.90-0.99), respectively. The area under ROC curve (AUC) was 0.927.

**Figure 4.**
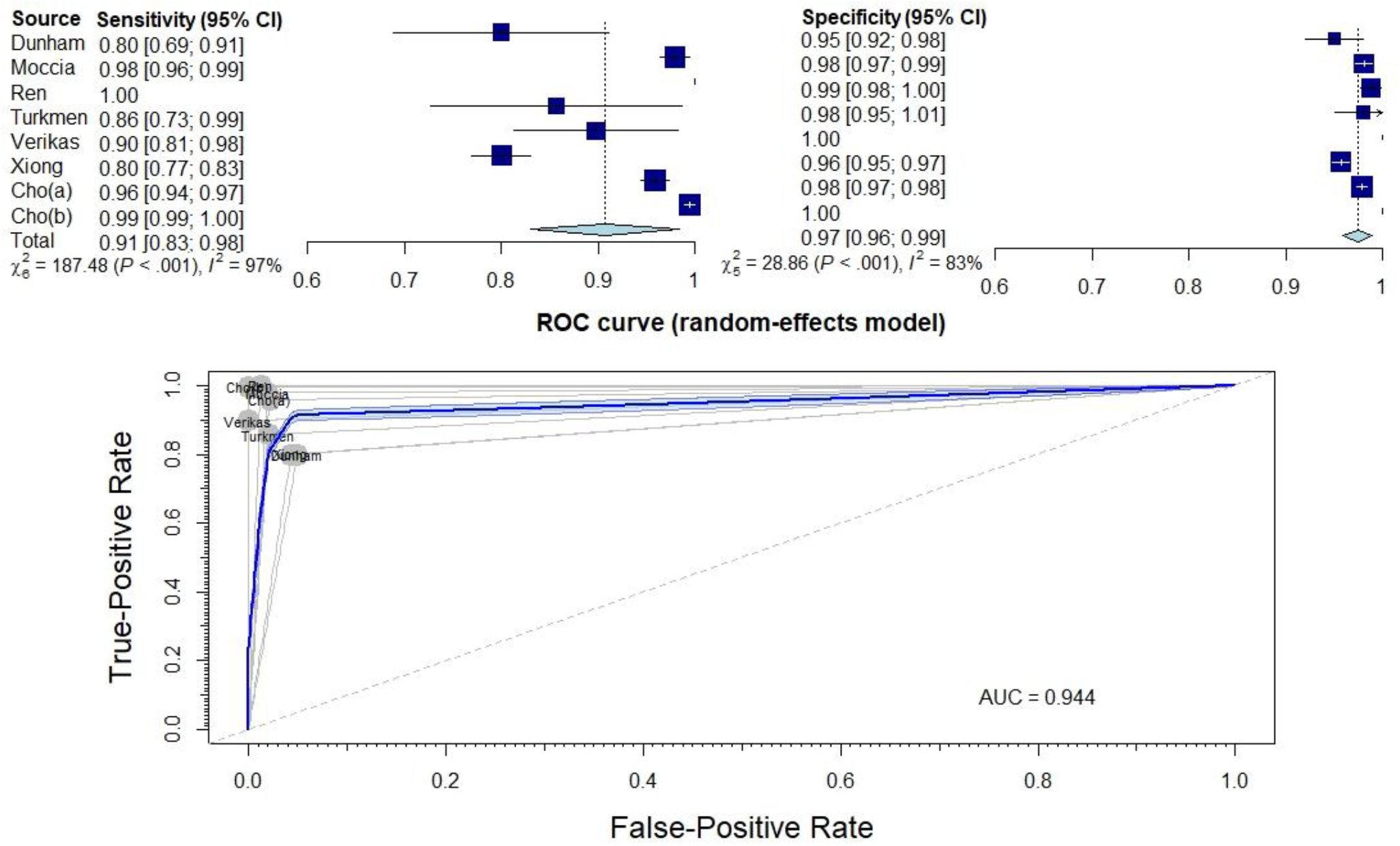
Forest plot and ROC curve illustrating the diagnostic performance of AI identifying healthy laryngeal tissue.

**Figure 5.**
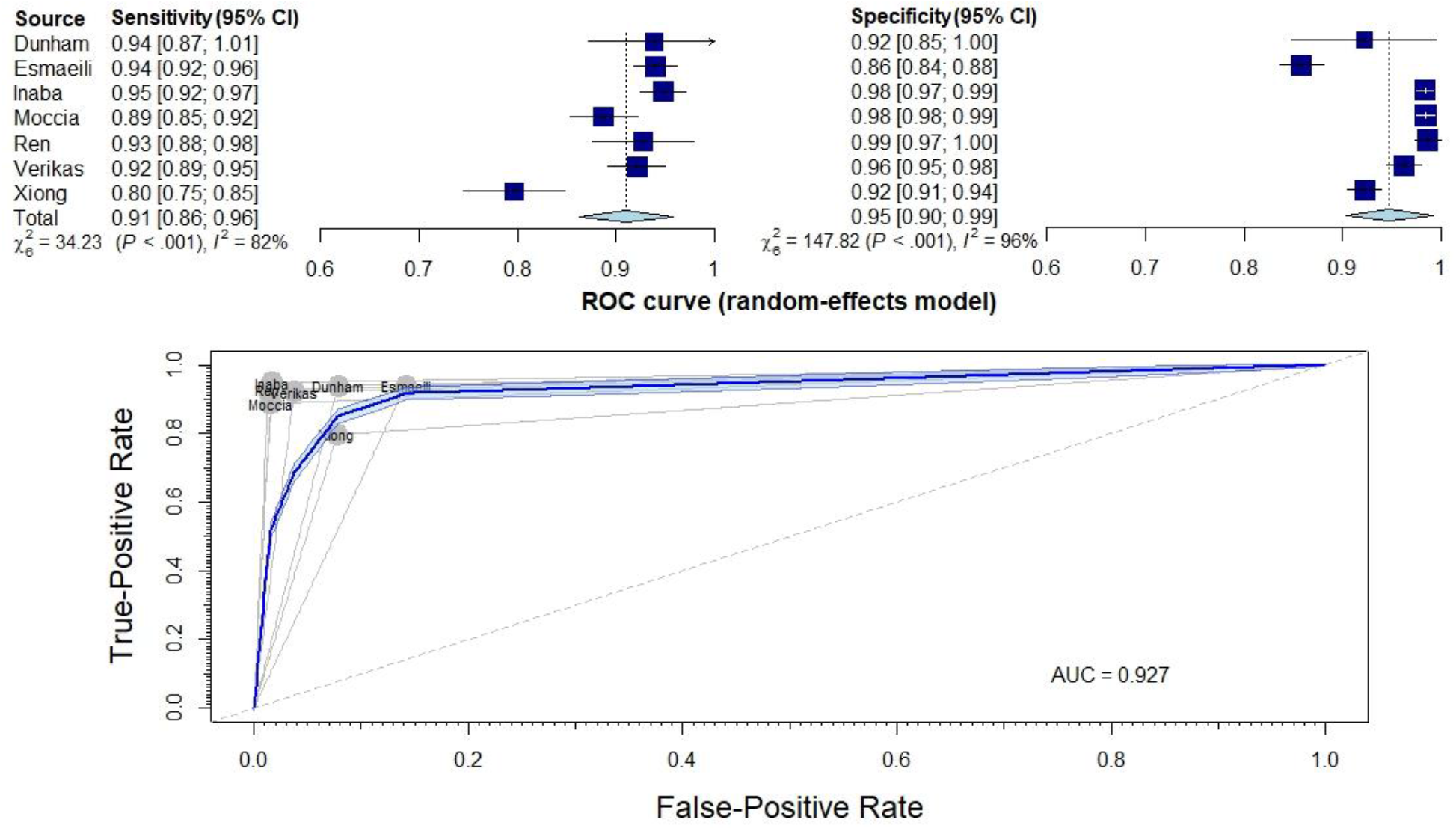
Forest plot and ROC curve illustrating the diagnostic performance of AI distinguishing benign and malignant laryngeal lesions.

**Figure 6.**
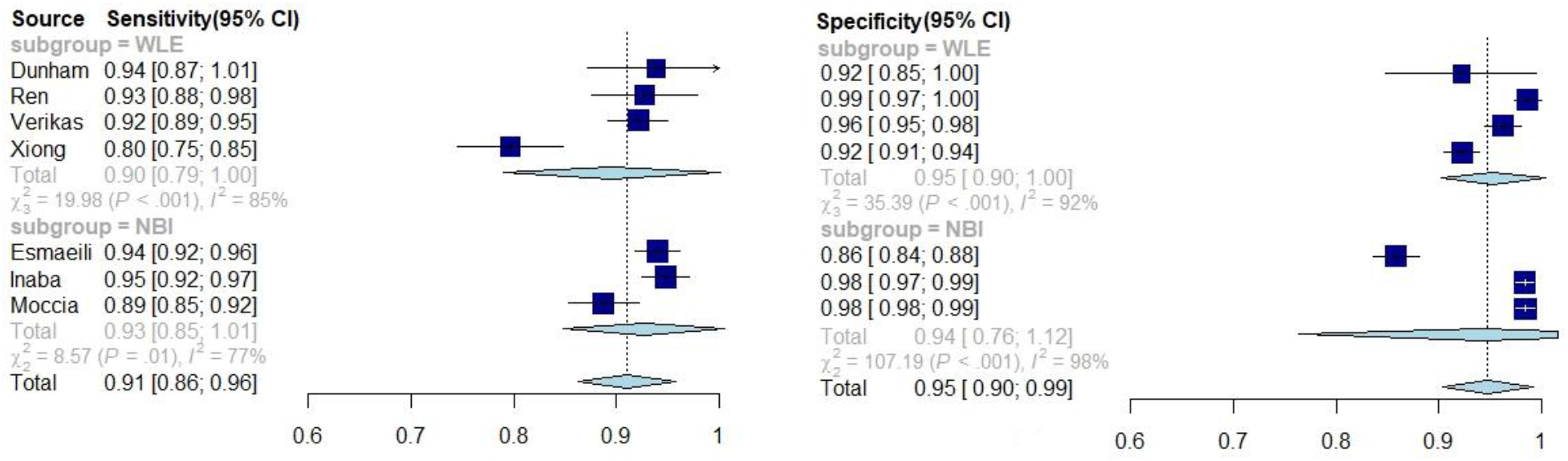
Forest plot illustrating the differences in diagnostic performance of AI using WLE and NBI.

The polled analysis revealed also a significant variation between studies (Sensitivity: τ^2^ = 0.0021 (95%CI: 0.0006-0.131), I^2^ = 82.5% (95%CI: 65.1%-91.2%); Specificity: τ^2^ = 0.0022 (95%CI: 0.0008-0.0109), I^2^ = 95.9% (95%CI: 93.6%-97.4%); p-value for both analysis <0.0001).

### 3.6. Comparison of diagnostics using WL and NBI

The last part of the analysis concerns the comparison of the results depending on the performed endoscopic method of WLE or NBI. This part of the analysis concerns the studies differentiating benign and malignant lesions in the larynx. The sensitivity of AI was higher for NBI (0.93, 95% CI: 0.85-1.01) than for WLE (0.90, 95% CI: 0.79-1.00). In turn, for specificity, the results were very similar, respectively 0.94 (95% CI: 0.76-1.12) for NBI and 0.95 (95% CI: 0.9-1.00) for WLE. The test for subgroup differences suggests that there is no statistically significant subgroup effect (p = 0.406 for sensitivity and p = 0.817 for specificity).

## 4. Discussion

### 4.1. Main findings

AI shows extremely high accuracy, sensitivity and specificity in assessing images of laryngeal lesions. The accuracy of cited studies differs between 0.806 to 0.997. Such high values indicate the great utility of AI in laryngology and provide potential opportunities to introduce AI into diagnostic standards. The regression model of accuracy of 9 included studies shows a statistically significant trend between accuracy of AI diagnoses and number of images (p = 0.0004). This means that the key element to improve the quality of AI models in the assessment of laryngeal lesions is the increase in the number of images used, while maintaining high-quality pre-processing method.

In the second part, the ability of AI to identify healthy tissue was assessed. The pooled sensitivity and specificity were 0.91 and 0.97, respectively, which indicates a very high efficiency. Depending on the study, healthy tissue was differentiated with malignant lesions, such as cancer or severe dysplasia [26, 27, 30, 32], but also with benign lesions, such as nodules, polyps, Reinke’s edemas, granulomas or vocal fold palsies [22, 23, 27, 28, 32]. Although it is difficult to indicate the clinical usefulness of AI on this basis, the results indicate its enormous potential, and it may help young doctors learn the correct diagnosis of laryngeal lesions, in particular, to differentiate benign lesions.

The next part evaluated the most important step in the diagnostics of laryngeal lesions, i.e. the differentiation of benign and malignant lesions. AI also performed very well - pooled sensitivity was 0.91 and pooled specificity was 0.95. Particularly high specificity indicates the ability of AI to discriminate patients with benign lesions from those with malignancies. These results confirm the high utility of AI in clinical practice.

The modern method of endoscopy, NBI, allows the enhanced visualization of vascular patterns and identification of neoangiogenesis accompanying carcinogenesis, which facilitates the differentiation of malignant lesions comparing with WLE in clinical practice [2, 6]. The results of AI accuracy for both methods were also confronted, and the results for AI assessment were comparable regardless the used technology, what is opposite to accuracy obtained with ENT specialists evaluation. Sensitivity and specificity of AI for both methods were 0.90 and 0.95 (for WLE) and 0.93 and 0.94 (for NBI), respectively. The analysis of subgroup differences shows that for AI there are no statistically significant differences in the accuracy of differentiating benign and malignant lesions in the WLE and NBI (p = 0.406 - 0.817).

### 4.2. Association with other studies

The application of AI in endoscopic evaluation is currently the subject of intense researches, especially in the digestive track endoscopy. The main task is to enhance the performance and resolve limitations related to experience and uncertainty, and therefore implement in modern instruments systems with automatic detection of pathologies. With regard to laryngeal endoscopy, the topic is still at its initial stage, however a significant increase in those researches has been observed in the last two years and the subject will be certainly intensively explored. At this early stage it is recommended to evaluate the main strategies of analysis and indicate importance of consistent data collection, homogeneity of nomenclature and comparable images amounts and technical aspects related to image processing.

For clinical reasons, the part of our meta-analysis focusing on distinguishing malignant and benign lesions seems to be the most crucial. The technical improvement of endoscopic images and their widespread in previous decade allowed more efficient preoperative diagnosis and therefore more accurate treatment strategy for those patient. There have been performed many studies assessing the effectiveness of these so-called “optical biopsy” in detecting malignant lesions. In the work of Davaris et al. [33] three experienced otorhinolaryngologists assessed endoscopic WLE and NBI images of laryngeal lesions, achieving sensitivity of 0.77 (95% CI: 0.688-0.853) and 0.933 (95% CI: 0.878-0.988) and specificity of 0.973 (95% CI: 0.956-0.991) and 0.973 (95% CI: 0.956-0.991), respectively. The reference standard was histopathologic examination. In the early meta-analysis of Zhou et al. from 2018 [34] summarizing 8 studies in the field of laryngeal lesions, the sensitivity and specificity in diagnosis of malignant lesions in NBI was 0.91 (95% CI: 0.885-0.931) and 0.915 (95% CI: 0.893-0.934), respectively. Later studies supported only the evidences with the vales of sensitivity and specificity ranging from 0.84 to 0.985 and from 0.889 to, 0.985, respectively [35-39]. It must be stressed, that in those researches the parameters of diagnostic accuracy were obtained based on evaluation by at least two specialists experienced with the method.

According to the presented results, the effectiveness of neural networks in the diagnosis of malignant changes does not differ from the assessments of professionals. The sensitivity and specificity were 0.91 (95% CI: 0.86-0.96) and 0.95 (95% CI: 0.90-0.99), respectively. The sensitivity was slightly higher when only the studies with the use of NBI light were assessed - 0.93 (95% CI: 0.85-1.01), which is consistent with the results of the studies cited above.

The results of the meta-analysis indicate that AI is a valuable tool for the assessment of laryngeal lesions and that the effectiveness of neural networks does not differ from the assessments of professionals. It would be particularly valuable to introduce AI in facilities that do not use NBI, because the sensitivity of the network assessments in white light (0.9) was higher than that of professionals (0.77), and the specificity was at a similar level (0.95 and 0.973, respectively).

However, it should be noted that one of the main goals of introducing AI tools in medicine is to support the work of young and inexperienced doctors. In the work of Nogués - Sabaté et al. [40] the effectiveness of the diagnosis of malignant lesions was compared using WLE and NBI images between experienced specialists and medical students. Inter-observer agreement among professionals was assessed both for WLE and NBI images as substantial (κ = 0.63 and 0.68, respectively), and for trainees as moderate (κ = 0.48 and 0.55). Żurek et al. [13] proved an increase in the sensitivity and specificity of the diagnosis of changes in the larynx along with the increase in the experience of the doctor assessing the images of laryngeal lesions (sensitivity 83.7% vs 98.1%, specificity 76.7% vs 80%). These results confirm the need to introduce additional diagnostic tools, especially for physicians with little experience.

### 4.3. Limitations

It should be pointed out to the inaccuracies in the results of the meta-analysis with the data provided in subchapter 4.2. First, the reference standard for the final diagnosis was varying in included studies and applied the histopathological result in major of them but also clinical diagnosis of an ENT specialist was accepted in studies analysing benign pathologies. Hence, the potential risk of misdiagnosis exist. Obviously the clinical differentiation and dysplasia staging between benign, premalignant or malignant lesions is burdened with an even greater risk of incorrect diagnosis what was proved in the researches [33, 34, 37]. Most of the analyzed studies, that involved malignant changes assessment with AI were referred to histopathological results. However, in the studies of Araújo et al. [20], Esmaeli et al. [24] and Moccia et al. [26] the diagnosis of ENT specialists was also accepted. On the other hand, in the works of Verikas et al. [29, 30] the reference standard has been specified as “clinical routine evaluation”. Therefore, the adopted reference standard may be subject to a priori error in these studies. For the future researches in the field the uniform referral norms should be accepted.

The other limitation of the meta-analysis is the considerable heterogeneity of the methodology, especially in terms of pre-processing of the images and the number of patients and images used. The smallest number of images used to assess laryngeal lesions was 120 [21] and the largest was 24,667 [27]. The pre-processing and methodology of the study is clearly related to the size of the study. A tendency was observed that the smaller the research sample, the more complicated pre-processing. In the smallest study of Barbalata et al. [21], the preparation of images for evaluation by AI was proceeded by many steps related to graphic processing of images, including specular reflections removal, ROI detection, blood vessel extraction, determination of vessel size. In contrast, the largest study, Ren et al. [27], did not process the images at all, but only manually removed duplicates and low-quality images. It should also be noted that when the studies with the same classification of laryngeal lesions (e.g. differentiation of benign and malignant lesions) are compared, the sensitivity and specificity of AI classification is at a similar level. This means that a complicated pre-processing method on a small number of images gives similar results to using a large image database. In the study by Verikas et al. [30] 785 images were used, and the AI distinguished benign and malignant lesions with a sensitivity and specificity of 92.12% and 96.26%, respectively. In contrast, in the study by Ren et al. [27] (24,667 images), the same classification achieved sensitivity and specificity of 92.78% and 98.66%, respectively.

The study by Arahujo et al. [20] used the first publicly available database of images of laryngeal lesions provided by Moccia et al. [41]. The Moccia et al. study [26] obtained an accuracy of 93%, while the same classification using a different methodology in the study by Arahujo et al. [20] obtained an accuracy of 98%.

Based on the above arguments, it should be concluded that the use of advanced methods of graphical image analysis on very large datasets will allow obtaining better results and increase clinical utility of AI models. This points to the need for large image databases and closer cooperation between medical and IT centres.

The standard in meta-analysis is the assessment of certainty of the evidence for outcomes. The limitation of this systematic review is the inability to use standard tools to assess the certainty of the evidence (like GRADE) [42], as there are still no standardized AI models, and each of the studies cited above used their own models. Although the results show great advantages and potential of AI, it is still not possible to recommend one specific tool for assessing laryngeal changes on this basis. As authors, we draw attention to the need of creation of publicly available databases of laryngeal lesions images and on this basis development of most accurate neural network model for laryngeal endoscopy. The development of researches on the clinical application of neural networks in this direction will allow for a comprehensive evaluation and will speed up identification of the most reliable tool in the diagnosis of laryngeal pathologies.

## 5. Conclusions

In assessing images of laryngeal lesions, AI demonstrates extraordinarily high accuracy, sensitivity, and specificity. Such high values indicate significant utility of AI and offer enhanced diagnostic tool in laryngology. The performance of AI diagnoses increases efficacy with the size of the image database used for learning and testing, and with the number of pre-processing steps involving extracting specific features of the images. The best way to increase the quality and utility of AI in diagnosis is to develop standards for evaluating images and to strengthen multicentre cooperation by sharing a database of images of laryngeal lesions, which allows building AI modes with the best performance, based on vast amount of images for learning and testing.

## Data Availability

All data produced in the present work are contained in the manuscript.

