## Supplementary Table 1 for "Artificial Intelligence in laryngeal endoscopy: Systematic Review and Meta-Analysis"

Table S1. Literature search strategy

| Search terms | neural network  OR  machine learning  OR  artificial intelligence | AND | larynx  OR  laryngeal  OR  vocal fold | AND | lesion  OR  benign  OR  malignant  OR  cancer  OR  carcinoma |
| --- | --- | --- | --- | --- | --- |
| Databases searched | PubMed, Embase, Cochrane, Scopus, Web of Science | | | | |
| Part of journals searched | Keywords in all parts of the articles (title, abstract and manuscript) | | | | |
| Years of search | All available till 15.10.2021 | | | | |
| Language | English | | | | |
| Types of studies to be included | Qualitative studies | | | | |
| Inclusion criteria | 1. Any clinical trial evaluating the application of neural network in endoscopic diagnosis of vocal fold lesions 2. The study concerns ENT patients with laryngeal lesions 3. The study evaluates at least one of the rates of neural network: accuracy, sensitive, specificity, true positive rate, false positive rate 4. No restriction regarding country, patient age, race, gender, publication language, and date | | | | |
| Exclusion criteria | 1. Study of neural-network in non-human subjects 2. Study with data not reliably extracted, duplicate, or overlapping 3. Abstract-only papers as preceding papers, conference, editorial, and author response theses and books 4. Articles without available full text available 5. Case reports, case series, and systematic review studies | | | | |
