## Supplementary Table 2 for "Artificial Intelligence in laryngeal endoscopy: Systematic Review and Meta-Analysis"

Table S2. The tailored QUADAS questions.

**DOMAIN 1: PATIENT SELECTION**

1. Was a consecutive or random sample of patients enrolled?
2. Was a case-control design avoided?
3. Did the study avoid inappropriate exclusions?
4. Has the classification been subjected to various lesions and not just a specific group, e.g. neoplastic changes and fragments of images?
5. Were images of different dimensions or whole images of the vocal folds used, and not just standardized image fragments of the same dimensions (which is associated with a lower utility of the network in clinical work)?

**DOMAIN 2: INDEX TEST(S)**

1. Were the index test results interpreted without knowledge of the results of the reference standard?
2. If a threshold was used, was it pre-specified?
3. Are the results clinically useful or was the aim of the study to improve the web from other articles with no relevance to its clinical utility?
4. Are there any cross-tables of the results of the artificial intelligence and not only the values of the rates (accuracy, sensitivity, specificity etc.)?
5. Was only the validation of the artificial intelligence performed without testing on another or additional set of images?

**DOMAIN 3: REFERENCE STANDARD**

1. Is the reference standard likely to correctly classify the target condition?
2. Were the reference standard results interpreted without knowledge of the results of the index test?
3. Was the diagnosis confirmed by histopathological examination when classifying malignant lesions?
4. Has the diagnosis been made by an ENT specialist with appropriate experience when classifying benign lesions, e.g. nodules, polyps?

**DOMAIN 4: FLOW AND TIMING**

1. Was there an appropriate interval between index test(s) and reference standard?
2. Did all patients receive a reference standard?
3. Did patients receive the same reference standard?
4. Were all patients included in the analysis?
5. Were all images classified in the same way (diagnosis of ENT specialist or histopathological examination)?
6. Were all images previously classified by the same ENT or histopathological examination?
7. Were all available images used without any exclusions?
