## Supplementary Table 3 for "Artificial Intelligence in laryngeal endoscopy: Systematic Review and Meta-Analysis"

Table S3. Summary table of the studies included in the meta-analysis.

| **Author** | **Year** | **Study design** | **No. Patients** | **No. Images (total)** | **Aims and outcomes** | **Preprocessing and methodology** | **CNN** | **Reference standard** | **Whole images** | **NBI** | **Terminology in article** | **Corresponding vascular pattern in Ni classification** | **No. Patients in the class** | **No. Images in the class** |
| --- | --- | --- | --- | --- | --- | --- | --- | --- | --- | --- | --- | --- | --- | --- |
| Ara ´ujo | 2019 | Retrospective | 33 | 1320 | Classification into four tissue classes: He (healthy tissue), Hbv (tissue with hypertrophic vessels), Le (tissue with leukoplakia) and IPCL (tissue with intrapapillary capillary loops) | Gaussian smoothing, texture-based global descriptors, first-order statistics and CNN-based learned features | 1 | ENT diagnose | 0 | 1 | He (healthy tissue) | I | - | 330 |
|  |  |  |  |  |  |  |  |  |  |  | Hbv (tissue with hypertrophic vessels) | II | - | 330 |
|  |  |  |  |  |  |  |  |  |  |  | Le (tissue with leukoplakia) | III | - | 330 |
|  |  |  |  |  |  |  |  |  |  |  | IPCL (tissue with intrapapillary capillary loops) | IV + V | - | 330 |
| Barbalata | 2016 | Retrospective | 60 | 120 | Classification into malignant versus benign lesions | Specular Reflections Removal, ROI detection, Blood Vessel Extraction, Classification based on vessel size | 0 | Histopathology | 1 | 1 | Benign lesions | I + II | 17 | 34 |
|  |  |  |  |  |  |  |  |  |  |  | Malignant lesions | IV + V | 43 | 86 |
| Cho | 2021 | Retrospective | 4106 | 4106 | Classification into normal larynx, cysts, nodules, polyps, leukoplakia, laryngeal papillomas, Reinke’s edema, granulomas and vocal cord palsies | Manually cropping vocal fold images to choose ROI, CNN with pretrained model | 1 | ENT diagnose | 1 | 0 | normal larynx | I | 742 | 742 |
|  |  |  |  |  |  |  |  |  |  |  | cysts | I + II | 347 | 347 |
|  |  |  |  |  |  |  |  |  |  |  | nodules | I | 171 | 171 |
|  |  |  |  |  |  |  |  |  |  |  | polyps | I + II | 948 | 948 |
|  |  |  |  |  |  |  |  |  |  |  | leukoplakia | III | 775 | 775 |
|  |  |  |  |  |  |  |  |  |  |  | laryngeal papillomas | IV+V | 120 | 120 |
|  |  |  |  |  |  |  |  |  |  |  | Reinke’s edema | I + II | 149 | 149 |
|  |  |  |  |  |  |  |  |  |  |  | granulomas | I + II | 296 | 296 |
|  |  |  |  |  |  |  |  |  |  |  | vocal cord palsies | I | 558 | 558 |
| Dunham | 2020 | Retrospective |  | 19353 | Binary classification into malignant– premalignant and benign lesions | Selection using a structural similarity index (SSIM), image data augmentation, CNN with pretrained model | 1 | Benign lesions - ENT diagnose; Malignant lesions - histopathology | 1 | 0 | **benign class** | I + II | - | 17423 |
|  |  |  |  |  |  |  |  |  |  |  | normals tissue | I | - | 3444 |
|  |  |  |  |  |  |  |  |  |  |  | nodules | I | - | 1816 |
|  |  |  |  |  |  |  |  |  |  |  | papillomas | IV + Va | - | 3633 |
|  |  |  |  |  | Classification of benign lesions: normal, nodules, papilloma, polyps and webs |  |  | ENT diagnose |  |  | polyps | I + II | - | 4577 |
|  |  |  |  |  |  |  |  |  |  |  | webs | I | - | 3953 |
|  |  |  |  |  |  |  |  |  |  |  | **malignant–premalignant class** | III + IV + V | - | 1930 |
|  |  |  |  |  |  |  |  |  |  |  | leukoplakia | III | - | 925 |
|  |  |  |  |  |  |  |  |  |  |  | carcinoma | IV+V | - | 1005 |
| Esmaeili | 2019 | Retrospective | 32 | 1485 | Classification of vascular patterns: “order”, “disorder” and “very disorder” | Homogenization of images, vessel enhancement with Frangi filter, skeletonization procedure of vessels, calculating five indicators (HGD, RIA, ANG, DIS, CUR) of vascular patterns, classification using SVM, k-nearest neighbors (kNN), and random forests (RF) | 0 | ENT diagnose | 0 | 1 | order vascular pattern | I | 32 | 1485 |
|  |  |  |  |  |  |  |  |  |  |  | disorder vascular pattern | II |  |  |
|  |  |  |  |  |  |  |  |  |  |  | very disorder vascular pattern | IV+V |  |  |
|  |  |  | 20 | 890 | Classification of benign histopathologies - cyst, polyp & reinke’s edema, papilloma, and dysplasia mild |  |  | Histopathology |  |  | **benign class** | I + II | 20 | 890 |
|  |  |  |  |  |  |  |  |  |  |  | cyst | I + II | 3 | 150 |
|  |  |  |  |  |  |  |  |  |  |  | polyp | I + II | 4 | 130 |
|  |  |  |  |  |  |  |  |  |  |  | reinke’s edema | I + II | 5 | 250 |
|  |  |  |  |  |  |  |  |  |  |  | papilloma | IV + Va | 5 | 230 |
|  |  |  |  |  |  |  |  |  |  |  | dysplasia mild | IV | 3 | 130 |
|  |  |  | 11 | 465 | Classification of malignant histopathologies: dysplasia severe, carcinoma in situ and carcinoma |  |  | Histopathology |  |  | **malignant histopathologies** | IV + V | 11 | 465 |
|  |  |  |  |  |  |  |  |  |  |  | dysplasia severe | IV + V | 4 | 130 |
|  |  |  |  |  |  |  |  |  |  |  | carcinoma in situ | IV + V | 4 | 155 |
|  |  |  |  |  |  |  |  |  |  |  | carcinoma | V | 3 | 180 |
|  |  |  | 31 | 1355 | Classification into benign and malignant histopathologies |  |  | Histopathology |  |  | - | - | 31 | 1355 |
| Inaba | 2020 | Retrospective | 374 | 2400 | Classification into superficial laryngopharyngeal cancer and normal tissue | Manually cropping vocal fold images to choose ROI, CNN with pretrained model | 1 | Histopathology | 1 | 1 | superficial laryngopharyngeal cancer (SLPC) | IV + V | 174 | 800 |
|  |  |  |  |  |  |  |  |  |  |  | normal laryngopharyngeal mucosa | I | 200 | 1600 |
| Moccia | 2017 | Retrospective | 33 | 1320 | Classification into: tissue with IPCL-like vessels, leukoplakia, tissue with hypertrophic vessels and healthy tissue | Anisotropic diffusion filtering, specular reflections masking, selecting of squared patches, extraction of texture-based global descriptors and first-order statistics, classification using SVM, k-nearest neighbors (kNN), naive Bayes (NB) and random forest (RF) | 0 | ENT diagnose | 0 | 1 | healthy tissue | I | - | 330 |
|  |  |  |  |  |  |  |  |  |  |  | tissue with hypertrophic vessels | II | - | 330 |
|  |  |  |  |  |  |  |  |  |  |  | leukoplakia | III | - | 330 |
|  |  |  |  |  |  |  |  |  |  |  | tissue with IPCL-like vessels | IV + V | - | 330 |
| Ren | 2020 | Retrospective | 9231 | 24667 | Classification into normal tissue, vocal nodule, polyps, leukoplakia and malignancy | Removing duplicated images, with low resolution, and without vocal cords, classification using CNN with pretrained model | 1 | Benign lesions - ENT diagnose; Malignant lesions - histopathology | 1 | 0 | normal tissue | I | - | 10215 |
|  |  |  |  |  |  |  |  |  |  |  | vocal nodule | I | - | 5807 |
|  |  |  |  |  |  |  |  |  |  |  | polyps | I+II | - | 2995 |
|  |  |  |  |  |  |  |  |  |  |  | leukoplakia | III | - | 2120 |
|  |  |  |  |  |  |  |  |  |  |  | malignancy | IV+V | - | 3530 |
| Turkmen | 2015 | Retrospective | 70 | 124 | Categorization into vocal folds into healthy, nodule, polyp, sulcus vocalis, and laryngitis classes | Detection of vocal folds based on Histogram of Oriented Gradients (HOG), segmentation of glottis, and normalization of vocal fold images; extraction of vocal fold edge and vessel features; classification using Naive Bayes, multilayer perceptron, k-nearest neighbors (KNN), support vector machine (SVM), and random forest (RF) | 0 | ENT diagnose | 0 | 0 | healthy tissue | I | 14 | 28 |
|  |  |  |  |  |  |  |  |  |  |  | nodule | I | 10 | 20 |
|  |  |  |  |  |  |  |  |  |  |  | polyp | I + II | 16 | 17 |
|  |  |  |  |  |  |  |  |  |  |  | sulcus vocalis | I | 15 | 29 |
|  |  |  |  |  |  |  |  |  |  |  | laryngitis | II | 15 | 30 |
| Verikas | 2007 | Retrospective |  | 785 | Clasification into nodular, diffuse, and healthy classes. | Extraction of colour, texture, and geometrical features; classification using SVM method with different kernel degrees | 0 | Clinical routine evaluation of patients | 1 | 0 | healthy tissue | I | - | 49 |
|  |  |  |  |  |  |  |  |  |  |  | nodular class (polyps, nodules, cysts) | I + II | - | 406 |
|  |  |  |  |  |  |  |  |  |  |  | diffuse class (papillomata, keratosis, carcinoma) | IV + V | - | 330 |
| Verikas | 2007 | Retrospective |  | 785 | Clasification into nodular, diffuse, and healthy classes. | Extraction of features: colour, texture, geometry, gradient and frequency contentusing Fourier spectrum; classification using SVM with the polynomial kernel of degree one to three | 0 | Clinical routine evaluation of patients | 1 | 0 | healthy tissue | I | - | 49 |
|  |  |  |  |  |  |  |  |  |  |  | nodular class (polyps, nodules, cysts) | I + II | - | 406 |
|  |  |  |  |  |  |  |  |  |  |  | diffuse class (papillomata, keratosis, carcinoma) | IV + V | - | 330 |
| Xiong | 2019 | Retrospective | 2208 | 14897 | Classification into Urgent versus Non-urgent subjects | Manually selecting images, CNN with pretrained model | 1 | Histopathology | 1 | 0 | **Non-urgent subjects** | I + II | 1712 | 10554 |
|  |  |  |  |  |  |  |  |  |  |  | **Urgent subjects** | IV + V | 496 | 4388 |
|  |  |  |  |  | Classification into laryngeal cancer (LCA), precancerous laryngeal lesions (PRELCA), benign laryngeal tumors (BLT) and normal tissues (NORM) |  |  |  |  |  | normal tissues (NORM) | I | 770 | 3602 |
|  |  |  |  |  |  |  |  |  |  |  | benign laryngeal tumors (BLT) | I+II | 942 | 6952 |
|  |  |  |  |  |  |  |  |  |  |  | precancerous laryngeal lesions (PRELCA) | III + IV + V | 246 | 1936 |
|  |  |  |  |  |  |  |  |  |  |  | laryngeal cancer (LCA) | V | 250 | 2452 |
| Cho | in press | Retrospective | 2216 | 2216 | Classification into normal and abnormal vocal fold tissue | Manually cropping vocal fold images to choose ROI, CNN with pretrained model | 1 | Histopathology | 1 | 0 | normal tissue | I | 899 | 899 |
|  |  |  |  |  |  |  |  |  |  |  | abnormal tissue | II + III + IV+ V | 1317 | 1317 |
