## Supplementary Table 4 for "Artificial Intelligence in laryngeal endoscopy: Systematic Review and Meta-Analysis"

Table S4. The results of the QUADAS-2 bias and applicability evaluation.

| **Study (Title/Author/Year)** | | | **RISK OF BIAS** | | | | **APPLICABILITY CONCERNS** | | |
| --- | --- | --- | --- | --- | --- | --- | --- | --- | --- |
|  |  |  | **PATIENT SELECTION** | **INDEX TEST** | **REFERENCE STANDARD** | **FLOW AND TIMING** | **PATIENT SELECTION** | **INDEX TEST** | **REFERENCE STANDARD** |
| Learned and handcrafted features for early-stage laryngeal SCC diagnosis | Ara ´ujo | 2019 | ☹ | ? | ☺ | ☺ | ☺ | ☺ | ☺ |
| Optical Biopsy: Automated Classification of Airway Endoscopic Findings Using a Convolutional Neural Network | Dunham | 2020 | ☺ | ☺ | ☺ | ☺ | ☺ | ☺ | ☺ |
| Novel automated vessel pattern characterization of larynx contact endoscopic video images | Esmaeili | 2019 | ? | ☺ | ☺ | ☺ | ☺ | ☺ | ☺ |
| Artificial intelligence system for detecting superficial laryngopharyngeal cancer with high efficiency of deep learning | Inaba | 2020 | ☺ | ☺ | ☺ | ☺ | ☺ | ☺ | ☺ |
| Confident texture-based laryngeal tissue classification for early stage diagnosis support | Moccia | 2017 | ☹ | ☺ | ☺ | ☺ | ☺ | ☺ | ☺ |
| Automatic Recognition of Laryngoscopic Images Using a Deep-Learning Technique | Ren | 2020 | ☺ | ☺ | ☺ | ☺ | ☺ | ☺ | ☺ |
| Classification of laryngeal disorders based on shape and vascular defects of vocal folds | Turkmen | 2015 | ? | ☹ | ? | ☺ | ☺ | ☺ | ☺ |
| A kernel-based approach to categorizing laryngeal images | Verikas | 2007 | ? | ☹ | ? | ? | ? | ☺ | ☺ |
| Multiple feature sets based categorization of laryngeal images | Verikas | 2007 | ? | ? | ? | ☺ | ? | ☺ | ☺ |
| Computer-aided diagnosis of laryngeal cancer via deep learning based on laryngoscopic images | Xiong | 2019 | ☺ | ☺ | ☺ | ☺ | ☺ | ☺ | ☺ |
| Diagnostic Accuracies of Laryngeal Diseases Using a Convolutional Neural Network-Based Image Classification System | Cho | 2021 | ☺ | ☺ | ☺ | ☺ | ☺ | ☺ | ☺ |
| Laryngeal Tumor Detection and Classification in Endoscopic Video | Barbalata | 2016 | ☺ | ? | ☺ | ☺ | ☺ | ☺ | ☺ |
| Comparison of Convolutional Neural Network Models for Determination of Vocal Fold Normality in Laryngoscopic Images | Cho | in press | ☺ | ? | ☺ | ☺ | ☺ | ☺ | ☺ |
