## Supplementary Table 5 for "Artificial Intelligence in laryngeal endoscopy: Systematic Review and Meta-Analysis"

Table S5. Raw data of the included studies.

TP – true positive; TN – true negative; FP – false positive; FN – false negative; SE – standard error; LCI – lower 95% confidence interval; UCI – upper 95% confidence interval

| **Author** | **Year** | **TP** | **TN** | **FP** | **FN** | **ACCURACY** | **SENSITIVITY** | | | | **SPECIFICITY** | | | |
| --- | --- | --- | --- | --- | --- | --- | --- | --- | --- | --- | --- | --- | --- | --- |
|  |  |  |  |  |  |  | **Value** | **SE** | **95% LCI** | **95% UCI** | **Value** | **SE** | **95% LCI** | **95% UCI** |
| **IDENTIFICATION OF HEALTHY LARYNGEAL TISSUE** | | | | | | | | | | | | | | |
| Dunham | 2020 | 40 | 190 | 10 | 10 | 0,92 | 0,80 | 0,06 | 0,69 | 0,91 | 0,95 | 0,02 | 0,92 | 0,98 |
| Moccia | 2017 | 323 | 971 | 19 | 7 | 0,98 | 0,98 | 0,01 | 0,96 | 0,99 | 0,98 | 0,00 | 0,97 | 0,99 |
| Ren | 2020 | 100 | 395 | 5 | 0 | 0,99 | 1,00 | 0,00 | 1,00 | 1,00 | 0,99 | 0,01 | 0,98 | 1,00 |
| Turkmen | 2015 | 24 | 94 | 2 | 4 | 0,95 | 0,86 | 0,07 | 0,73 | 0,99 | 0,98 | 0,01 | 0,95 | 1,01 |
| Verikas | 2007 | 44 | 736 | 0 | 5 | 0,99 | 0,90 | 0,04 | 0,81 | 0,98 | 1,00 | 0,00 | 1,00 | 1,00 |
| Xiong | 2019 | 528 | 2075 | 94 | 132 | 0,92 | 0,80 | 0,02 | 0,77 | 0,83 | 0,96 | 0,00 | 0,95 | 0,97 |
| Cho | 2021 | 694 | 3309 | 73 | 30 | 0,97 | 0,96 | 0,01 | 0,94 | 0,97 | 0,98 | 0,00 | 0,97 | 0,98 |
| Cho | in press | 894 | 1317 | 0 | 5 | 1,00 | 0,99 | 0,00 | 0,99 | 1,00 | 1,00 | 0,00 | 1,00 | 1,00 |
| **DIFFERENTIATION BETWEEN BENING AND MALIGNANT LESIONS** | | | | | | | | | | | | | | |
| Dunham | 2020 | 46 | 47 | 4 | 3 | 0,93 | 0,94 | 0,03 | 0,87 | 1,01 | 0,92 | 0,04 | 0,85 | 1,00 |
| Esmaeili | 2019 | 437 | 764 | 126 | 28 | 0,89 | 0,94 | 0,01 | 0,92 | 0,96 | 0,86 | 0,01 | 0,84 | 0,88 |
| Inaba | 2020 | 328 | 787 | 13 | 18 | 0,97 | 0,95 | 0,01 | 0,92 | 0,97 | 0,98 | 0,00 | 0,97 | 0,99 |
| Moccia | 2017 | 284 | 636 | 10 | 36 | 0,95 | 0,89 | 0,02 | 0,85 | 0,92 | 0,98 | 0,00 | 0,98 | 0,99 |
| Ren | 2020 | 90 | 294 | 4 | 7 | 0,97 | 0,93 | 0,03 | 0,88 | 0,98 | 0,99 | 0,01 | 0,97 | 1,00 |
| Verikas | 2007 | 304 | 438 | 17 | 26 | 0,95 | 0,92 | 0,01 | 0,89 | 0,95 | 0,96 | 0,01 | 0,95 | 0,98 |
| Xiong | 2019 | 188 | 867 | 73 | 48 | 0,90 | 0,80 | 0,03 | 0,75 | 0,85 | 0,92 | 0,01 | 0,91 | 0,94 |
| **IDENTIFICATION OF LEUKOPLAKIA** | | | | | | | | | | | | | | |
| Moccia | 2017 | 304 | 956 | 24 | 26 | 0,96 | 0,92 | 0,01 | 0,89 | 0,95 | 0,98 | 0,00 | 0,97 | 0,99 |
| Ren | 2020 | 91 | 395 | 5 | 9 | 0,97 | 0,91 | 0,03 | 0,85 | 0,97 | 0,99 | 0,01 | 0,98 | 1,00 |
| Cho | 2021 | 690 | 3300 | 84 | 32 | 0,97 | 0,96 | 0,01 | 0,94 | 0,97 | 0,98 | 0,00 | 0,97 | 0,98 |
